# From Housing to Hotspots: Integrating a Housing-Based Measure of Individual Socioeconomic Status with Geospatial Analysis to Target Colorectal Cancer Screening in Rural Communities

**DOI:** 10.64898/2026.08.28.26361444

**Authors:** Rebecca Yao, Chung-Il Wi, Madison J. Beenken, Dave Watson, Philip H. Wheeler, Mike Finch, Dan P. Kelleher, Gokhan Anil, Trent Anderson, Kathy A. Madden, Scott H. Okuno, Folakemi T. Odedina, Erin C. Westfall, Eunice Y. Park, Pravesh Sharma, Sagar B. Dugani, Randy M. Foss, Brandon Hidaka, Jessica L. Sosso, Shivani Sabarish, Gurpreet Singh, Nahyr Lugo-Fagundo, James F. Howick V, W. Ray Kim, Andrew D. Calvin, Cheryl L. Walker-McGill, Lior Rennert, Young J. Juhn, James R. Cerhan, Brian A. Lynch

## Abstract

**Purpose:** This study assesses the association between colorectal cancer (CRC) screening and a validated, housing-based measure of individual-level socioeconomic status (SES, called HOUSES hereafter) within rural communities and determines whether HOUSES-integrated geospatial analysis can be used to tailor interventions.

**Methods:** We used CRC screening data from a subset of Mayo Clinic Midwest patients living in cities without ready access to routine care in the Mayo Clinic Health System in 2019 to represent rural communities. At the individual level, we assessed the association between CRC screening rates and the HOUSES index, adjusting for age, sex, race/ethnicity, comorbidity, distance from home address to clinic, and area deprivation index, using a multilevel mixed-effects logistic regression model. Additionally, we conducted geospatial analysis to examine the correlation between hotspots of 1) lower CRC screening rates and 2) lower SES of the subject population (HOUSES quartile 1).

**Findings:** Among 34,489 individuals (median age 64.0 years, 52.4% female), those with the lowest SES (HOUSES Q1) had 37% lower odds of being CRC screening adherent than those with the highest SES (HOUSES Q4) (adj. OR [95% CI]: 0.63 [0.58-0.69]). In the 14 identified HOUSES Q1 hotspots, there was a significant correlation in counts of HOUSES Q1 and low CRC screening (correlation coefficient=0.81).

**Conclusion:** Lower SES was significantly associated with lower CRC screening among rural populations. HOUSES-enabled geospatial analysis identified geographic hotspots with lower CRC screening rates for targeted interventions to address disparities in CRC screening in rural communities. HOUSES may be a useful digital tool for cancer preventive care and research.

## INTRODUCTION

Colorectal cancer (CRC) is the second leading cause of cancer-related deaths in the United States among men and women combined, with an estimated 52,900 deaths in 2025 [1]. Guideline-directed screening is a well-established measure for prevention and is widely credited as a key reason for declining CRC mortality over recent decades [2, 3]. However, CRC screening participation varies significantly among different population subgroups, with disparities by race, ethnicity, screen-eligible age, and residential setting (i.e., rural vs. urban) [4–7]. A systematic review found that rural residents were significantly less likely than urban residents to report ever completing CRC screening (pooled odds ratio = 0.81, 0.76-0.86) [5]. However, less is known about the extent to which these rural disparities in CRC screening are caused by individual social determinants of health (SDOH) [8]. This limits the development of targeted interventions, despite the well-recognized role of SDOH in shaping health outcomes [9, 10].

Socioeconomic status (SES), a key SDOH, is consistently associated with CRC screening participation, treatment adherence, and survival [11]. Although both individual- and neighborhood-level SES indicators contribute to lower CRC screening adherence [7, 11–13], individual-level SES is particularly important for designing patient-centered, targeted interventions. However, accurately capturing individual SES remains challenging, as it is often missing in commonly used datasets like electronic health records (EHRs) [14–16]. This gap may be especially consequential in rural settings, where limited local health system infrastructure can restrict screening access [5]. Because inequities often result from both differences in access to health-care resources and the ability to engage in preventive behaviors [17], the workload-capacity imbalance framework suggests that individuals at highest risk simultaneously face greater health care needs (e.g., delayed or missed CRC screening) and fewer resources or limited capacity to meet those demands [18, 19]. SES is one indicator of this capacity, reflecting an individual’s access to respond to those health needs by capturing access to material, social, and health-related resources. However, when individual-level SES is unavailable in EHRs, population-level interventions may fail to reach those most in need, a gap that is likely amplified in rural settings where limited local health system infrastructure can compound individual-level resource constraints [17, 18].

To address this gap, the HOUsing-based SocioEconomic Status (HOUSES) index is an individual-level SES that has been proven to be linked to a range of health outcomes, behaviors, health care access, and quality of care in adults and children [20–27]. By linking an individual’s home address to publicly available property data, the HOUSES index serves as a proxy for individual-level capacity to access health care and preventive services. Because HOUSES index relies on standardized county assessor data across the 50 states of the U.S., it is applicable in both urban and rural settings [21, 26, 28].

Rurality itself is commonly classified using administrative definitions such as Rural-Urban Commuting Area (RUCA) codes, based on population density and commuting patterns. However, as with aggregate SES measures, these classifications may not fully capture a community’s health system infrastructure, particularly in hub-and-spoke regional care models where care can vary locally regardless of standard rurality metrics. Thus, defining rurality based on proximity to local health-system infrastructure, rather than on administrative classification alone, may offer a more actionable lens for identifying communities where outreach or non-facility-based interventions could improve access to preventive care.

As a geocoded individual-level measure, HOUSES index integrates naturally with geographic information systems (GIS). GIS methods with heatmap generation have been used to visualize and address disparities in preventive cancer screening by mapping patterns of utilization, incidence, mortality, and health care access [29]. Such methods highlight areas with higher health needs based on unmet screening density. However, they may overlook individuals’ limited capacity to access resources and care, especially when health care resources are scarce. In rural areas, lower population density further weakens the utility of aggregate SES measures (e.g., area deprivation or social vulnerability indices) [30–32]. Integrating individual-level SES measures into geospatial analyses may, therefore, improve identification of populations with both high health needs and limited capacity to access resources and care. Further, the HOUSES index facilitates this geospatial analysis by integrating data from multiple sources, supporting spatial statistics, allowing multilevel analysis, and producing high-quality cartographic products and maps [26, 28, 33, 34].

Given the need to allocate limited preventive health resources strategically in rural communities, we hypothesize that the HOUSES index and related geospatial analysis can identify individuals at higher risk of CRC screening non-adherence in rural areas of Southern Minnesota, as well as communities characterized by both “higher health needs” and “limited capacity” due to lower SES defined by the HOUSES index.

## METHODS

### Study Setting and Population

Mayo Clinic Health System (MCHS) operates as a hub-and-spoke model with 16 hospitals and over 35 clinics as the spokes. While 15-20% of all Americans live in rural areas [17], about 47% of the population (382,087/806,661) and 53% of Census tracts (157 of 295 in the MCHS catchment area are considered rural) [35]. The statewide (MN, the current study setting) average CRC screening rate was 73.7% in 2020, slightly higher than the national average of 70.4% in 2020 [36, 37].

For the purposes of this study, we operationalized the rural analytic population as patients residing in communities within the MCHS catchment area that did not have an MCHS fixed primary care clinic location during the measurement year. This definition was selected to align with the study’s implementation objective: identifying geographically concentrated populations for whom community-based outreach, mobile health services, or other non-facility-based interventions may be most relevant. Although rurality can be defined using multiple county- or census tract-level classification systems, including RUCA codes, these administrative definitions may not fully capture local access to health system infrastructure in a hub-and-spoke regional care model. Therefore, our operational definition emphasizes the absence of local MCHS primary care infrastructure as a pragmatic marker of rural service context and potential outreach need. Patients residing in communities with an MCHS fixed primary care clinic location were excluded from the analytic cohort.

### Study Design

This is a retrospective cross-sectional study assessing CRC screening rates from January 1, 2019, to December 31, 2019, among adults aged 50-75 years. CRC screening adherence was defined according to Minnesota Community Measurement (MNCM) specifications, a statewide independent nonprofit organization that publicly reports standardized health care quality metrics across Minnesota health systems and provider organizations in MN [37]. MNCM measures are widely used for benchmarking preventive care quality and population health performance across health care organizations in Minnesota.

We utilized a subset of Mayo Clinic Midwest patients included in the MNCM 2020 reporting cycle (measurement year 2019). To focus on rural populations with limited access to health care services, we included patients residing in communities (cities) without a fixed primary care clinic of MCHS where patients must travel to receive in-person medical services. This operational definition was used as a surrogate marker for rural health care access and care delivery limitations. Rural communities were selected because rural populations experience persistently lower CRC screening rates, greater barriers to preventive care access, and higher socioeconomic disadvantage compared with urban populations, contributing to disparities in CRC outcomes. The study aimed to assess the association between CRC screening adherence and individual-level SES measured by the HOUSES index and to determine whether integrating HOUSES with geospatial analysis could help identify high risk areas within rural communities where targeted interventions may be most beneficial. This study was approved by the Institutional Review Board at Mayo Clinic (IRB # 19-009328).

### Definition of CRC screening (primary outcome)

We defined appropriate guideline-directed CRC screening as including the following [38–41]:

- Colonoscopy during the measurement period or the nine years prior,
- Flexible sigmoidoscopy during the measurement year or the four years prior,
- CT colonography during the measurement year or the four years prior,
- Fecal immunochemical test (FIT)-DNA during the measurement year or the two years prior, or
- Guaiac-based fecal occult blood test (gFOBT) or FIT during the measurement year

Individuals who met any of these five criteria were considered CRC screening adherent. It is noted that the recommended age for initiating CRC screening was lowered to 45 years after this study period, and this study evaluated screening adherence among adults aged 50-75 years old, based on the previous guideline range for average-risk individuals [38, 40].

### HOUsing-based SocioEconomic Status (HOUSES) Index as an individual-level SES measure

The HOUSES index was developed to address the lack of individual-level SES measures readily available in commonly used datasets such as EHRs [20, 27]. A z-score was calculated for four housing characteristics (square footage of the housing unit, estimated building value, number of bedrooms, number of bathrooms). These measures were standardized at the county level to reflect an individual’s SES relative to others within the same county and combined into a composite HOUSES z-score. The composite score was categorized into county-specific quartiles, with the lowest quartile (Q1) representing the lowest relative SES and the highest quartile (Q4) representing the highest relative SES within the county. The HOUSES index measures socioeconomic status using objectively assessed housing characteristics, reflecting household wealth and material resources beyond income alone [21, 26, 28, 33, 34]. Individuals’ addresses were geocoded and linked to publicly available property data from the local government assessors’ offices. To support dissemination and implementation, the HIPAA-compliant HOUSES Platform, which leverages cloud technology, was developed, delivering HOUSES as a nationwide and scalable SES index to support assessment of the generalizability of study findings like ours.

### Other Variables

Other variables were obtained from the EHR, which included demographic information such as age at baseline, sex assigned at birth, and self-identified race and ethnicity. Comorbidities were assessed using the Charlson Comorbidity Score, derived from the International Classification of Diseases - Clinical Modification 10 (ICD-10-CM) codes. The Area Deprivation Index (ADI), an area-level SES measure, is derived using data from the American Community Survey and incorporates 17 indicators of socioeconomic disadvantage, such as income, education, employment, and housing quality, at the census block group level [42, 43]. Each geographic area receives a composite ADI score, which is then ranked and standardized at the national level, with higher scores indicating greater socioeconomic deprivation. ADI for the individuals in this study was formulated by geocoding their addresses and matching to the existing dataset of ADI. Distance in miles from patient residence to the closest fixed primary care clinic of MCHS (spoke clinic) was calculated using the latitude and longitude points. All data were de-identified before analysis.

### Geospatial Analyses

After the addresses of all eligible individuals were geocoded using parcel-based geocoding methods, we employed the Environmental Systems Research Institute (ESRI) ArcMap 10.8 geographic information systems (GIS) software to perform geospatial analyses of the eligible individuals, using HOUSES index data and focusing on concentrations of individuals in the lowest quartile (Q1), and concentrations of non-adherent CRC screening cases outside the cities with a fixed primary care clinic of MCHS. Heatmaps were produced using raster-based kernel density estimation with a bandwidth of one mile and a cell size of 0.2 miles on each side (roughly equivalent to the area of four city blocks) to illustrate the spatial distribution of the subject population, CRC screening non-adherence, and HOUSES Q1 across the study region. As with past studies, we defined hotspots based on significant differences between observed and expected kernel densities [26, 28, 33, 34], where significance was determined by observed counts exceeding expected counts in a cluster of cells with a Poisson value of at least 95% confidence. We recognized in this study that targeted interventions should serve areas that reflect non-adherent individuals of efficiently served density and of significance in terms of relative difference [44]. In other words, hotspots should reflect proportions of need, and not areas where the number of individuals is a function chiefly of population size.

Based on this reasoning, potential hotspots were initially identified based on the density per square mile of HOUSES Q1 and CRC screening non-adherent individuals outside cities with a fixed primary care clinic of MCHS. Individuals who resided within the overlap of a HOUSES Q1 hotspot and low CRC screening hotspots were identified, and their characterization was compared with those living outside the overlapped hotspots. Geocoding errors encountered were addressed by excluding records with zero HOUSES values and by verification with Google Earth imagery.

### Data Analysis

Descriptive statistics and frequency distributions were generated for patient demographics and SES measures. Continuous measures were presented as median and quartiles, and categorical measures were presented as counts and percentages. Comparisons were made between patients who were CRC screening adherent versus those who were not, as well as those who resided within the overlap of a HOUSES Q1 hotspot and CRC lack of screening hotspot versus all others, using Chi-square tests for categorical variables and Kruskal-Wallis rank sum test for continuous variables. Hierarchical logistic regression modeling from the *lme4* package was used to evaluate the association between HOUSES quartiles and CRC screening [45]. The model was adjusted for age, sex, race/ethnicity, comorbidity, distance to spoke clinic, and ADI. The ADI was stratified into quartiles, with Q1 representing the highest SES and Q4 the lowest SES. To account for HOUSES being standardized within the county and the ADI being calculated at the census block group level, the model included county and census block group as nested random effects. Patients were seen at multiple locations across the region, so the clinic location was also included as a non-nested random effect. The significance level was set at 5%. All analyses were conducted using R version 4.4.1 (R Core Team (2024)).

## RESULTS

### Baseline characteristics

Out of the 79,988 individuals whose data were reported for MNCM 2020, 45,499 (56.9 %) individuals who resided in cities with a fixed primary care clinic of MCHS were excluded. A total of 34,489 eligible individuals were included in the study with a median age (interquartile range) of 64 (58-69), 52.4% female, 96.5% non-Hispanic White, and 14.0% HOUSES Q1 (Table 1). Overall, 27,316 (79.2%) were adherent to CRC screening. Compared with adherent individuals, those who were non-adherent to CRC screening guidelines were slightly more likely to be male (49.6% vs 47.1%), somewhat younger (62 years vs 64 years), Hispanic (1.6% vs 0.6%), non-White (5.0% vs 2.4%), greater distance to nearest spoke clinic (median 72.4mi vs. 64.6mi), and belonged to lower SES at the individual (HOUSES Q1: 18.4% vs. 12.9%) as well as the neighborhood (ADI Q4 12.1% vs. 8.7%) levels. Supplementary Figure 1 shows a bar plot of counts (%) of patients in different hotspots and corresponding screening status.

**Table 1.** Baseline characteristics of study individuals by CRC screening adherence status.

|  |  |  |  |  |
| --- | --- | --- | --- | --- |
|  | **CRC screening**<br>**adherent (n=27,316)** | **CRC screening non-**<br>**adherent (n=7,173)** | **Total**<br>**(N=34,489)** | **P-value** |
| **Age, years, median (Q1, Q3)** | 64 (59, 69) | 62 (56, 68) | 64 (58, 69) | <0.001<sup>1</sup> |
| **Female, n (%)** | 14,461 (52.9%) | 3,614 (50.4%) | 18,075 (52.4%) | <0.001<sup>2</sup> |
| **Race, n (%)** |  |  |  | <0.001<sup>2</sup> |
| White | 26,651 (97.6%) | 6,808 (95.0%) | 33,459 (97.1%) | <0.001<sup>2</sup> |
| American Indian/Alaska<br>Native | 39 (0.1%) | 9 (0.1%) | 48 (0.1%) |  |
| Asian | 137 (0.5%) | 60 (0.8%) | 197 (0.6%) |  |
| Black or African<br>American | 78 (0.3%) | 43 (0.6%) | 121 (0.4%) |  |
| Native Hawaiian/Pacific<br>Islander | 12 (<0.1%) | 3 (<0.1%) | 15 (<0.1%) |  |
| Other/Unknown | 392 (1.4%) | 241 (3.4%) | 633 (1.8%) |  |
| N-Miss | 7 | 9 | 16 |  |
| **Hispanic or Latino, n (%)** | 171 (0.6%) | 112 (1.6%) | 283 (0.8%) | <0.001<sup>2</sup> |
| **Charlson Comorbidity Index,**<br>**median (Q1, Q3)** | 2.0 (0.0, 3.0) | 2.0 (1.0, 2.0) | 2.0 (0.0, 3.0) | 0.703<sup>1</sup> |
| **Distance to Clinic (mi),**<br>**Median (Q1, Q3)** | 64.6 (32.6, 107.7) | 72.4 (41.7, 112.4) | 64.7 (34.5, 109.5) | <0.001<sup>1</sup> |
| **HOUSES Index Quartile, n (%)** |  |  |  | <0.001<sup>2</sup> |
| 4 (highest SES) | 9,673 (39.6%) | 2,058 (32.8%) | 11,731 (38.2%) |  |
| 3 | 6,910 (28.3%) | 1,696 (27.0%) | 8,606 (28.1%) |  |
| 2 | 4,678 (19.2%) | 1,362 (21.7%) | 6,040 (19.7%) |  |
| 1 (lowest SES) | 3,143 (12.9%) | 1,155 (18.4%) | 4,298 (14.0%) |  |
| N-Miss | 2912 | 902 | 3,814 |  |
| **Area Deprivation Index (ADI) in**<br>**quartile, n (%)** |  |  |  | <0.001<sup>2</sup> |
| 1 (highest SES) | 2,210 (9.1%) | 397 (6.3%) | 2,607 (8.5%) |  |
| 2 | 12,583 (51.6%) | 2,948 (47.1%) | 15,531 (50.7%) |  |
| 3 | 7,468 (30.6%) | 2,161 (34.5%) | 9,629 (31.4%) |  |
| 4 (lowest SES) | 2,111 (8.7%) | 756 (12.1%) | 2,867 (9.4%) |  |
| N-Miss | 2,944 | 911 | 3,855 |  |
| 1. Kruskal-Wallis rank sum test; 2. Pearson's Chi-squared test |  |  |  |  |

### Association of CRC screening with individual-level SES (HOUSES index)

Table 2 shows that CRC screening was significantly associated with HOUSES index after adjusting for demographic characteristics (age, sex, race/ethnicity), comorbidity, and ADI by using a multilevel mixed effects logistic regression model. Individuals with lower SES (HOUSES Q1) had significantly lower CRC screening compared to those with the highest SES (Q4) in a dose-response manner (adj. OR [95% CI]: 0.63 [0.58, 0.69] for Q1, 0.76 [0.70, 0.82] for Q2, and 0.87 [0.81. 0.94] for Q3). Living in higher-deprived areas (as defined by higher ADI), younger age, male, other race/ethnicity (vs. non-Hispanic white) were also independently associated with lower CRC screening rate.

**Table 2.** Multivariate analysis of association between HOUSES index and CRC screening rate (adj. Odds Ratio and 95% Confidence Interval)

|  |  |  |
| --- | --- | --- |
| **Term** | **Adjusted ORs (95% CI)** | **P-value** |
| **HOUSES index in quartile** |  |  |
| Q1 | 0.63 (0.58, 0.69) | <0.001 |
| Q2 | 0.76 (0.70, 0.82) | <0.001 |
| Q3 | 0.87 (0.81, 0.94) | <0.001 |
| Q4 (highest SES) | Ref | Ref |
| **Area Deprivation Index (ADI) in quartile** |  |  |
| Q1 (highest SES) | Ref | Ref |
| Q2 | 0.94 (0.83, 1.06) | 0.319 |
| Q3 | 0.82 (0.72, 0.94) | 0.005 |
| Q4 | 0.75 (0.63, 0.88) | <0.001 |
| **Age (per 1 year)** | 1.04 (1.03, 1.04) | <0.001 |
| **Sex** |  |  |
| Male | 0.89 (0.84, 0.94) | <0.001 |
| Female | Ref | Ref |
| **Race/Ethnicity** |  |  |
| Non-Hispanic White | Ref | Ref |
| Other | 0.49 (0.42, 0.56) | <0.001 |
| **Charlson Comorbidity Index (per 1 comorbidity)** | 1.02 (1.00, 1.03) | 0.074 |
| **Distance to clinic (mi)** | 1.00 (1.00, 1.00) | 0.803 |

#### Geospatial Comparison of Population Characteristics in Hotspot vs. Non-Hotspot Areas

There were 20 potential hotspots having a density of at least one HOUSES Q1 subject per cell (equivalent to 25 individuals per square mile) and 18 potential hotspots having at least one CRC-screening non-adherent subject per cell. Fourteen HOUSES Q1 hotspots showed a statistically significant association with higher-than-expected HOUSES Q1 concentrations. Of these 14 hotspots, 13 partially or completely overlapped with areas identified as significant CRC screening hotspots. Figures 1 and 2 display the geographic locations of the HOUSES Q1 and CRC hotspots, respectively. Figure 3 highlights the spatial overlap between HOUSES Q1 and CRC hotspots (correlation coefficient: 0.81).

**Figure 1.**
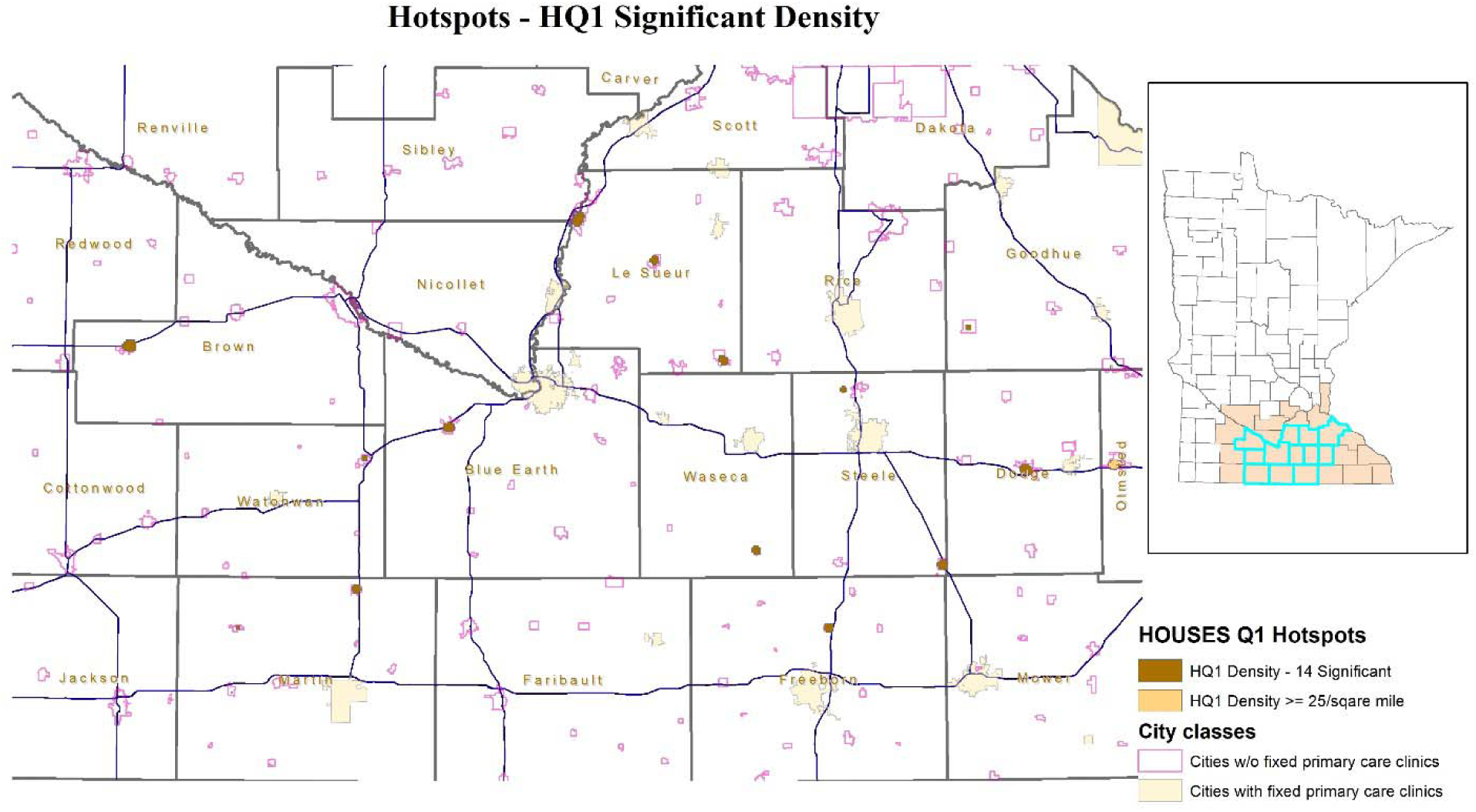
Geospatial Distribution of Low Socioeconomic Status as Measured by the HOUSES index (Quartile 1) in Rural Communities

**Figure 2.**
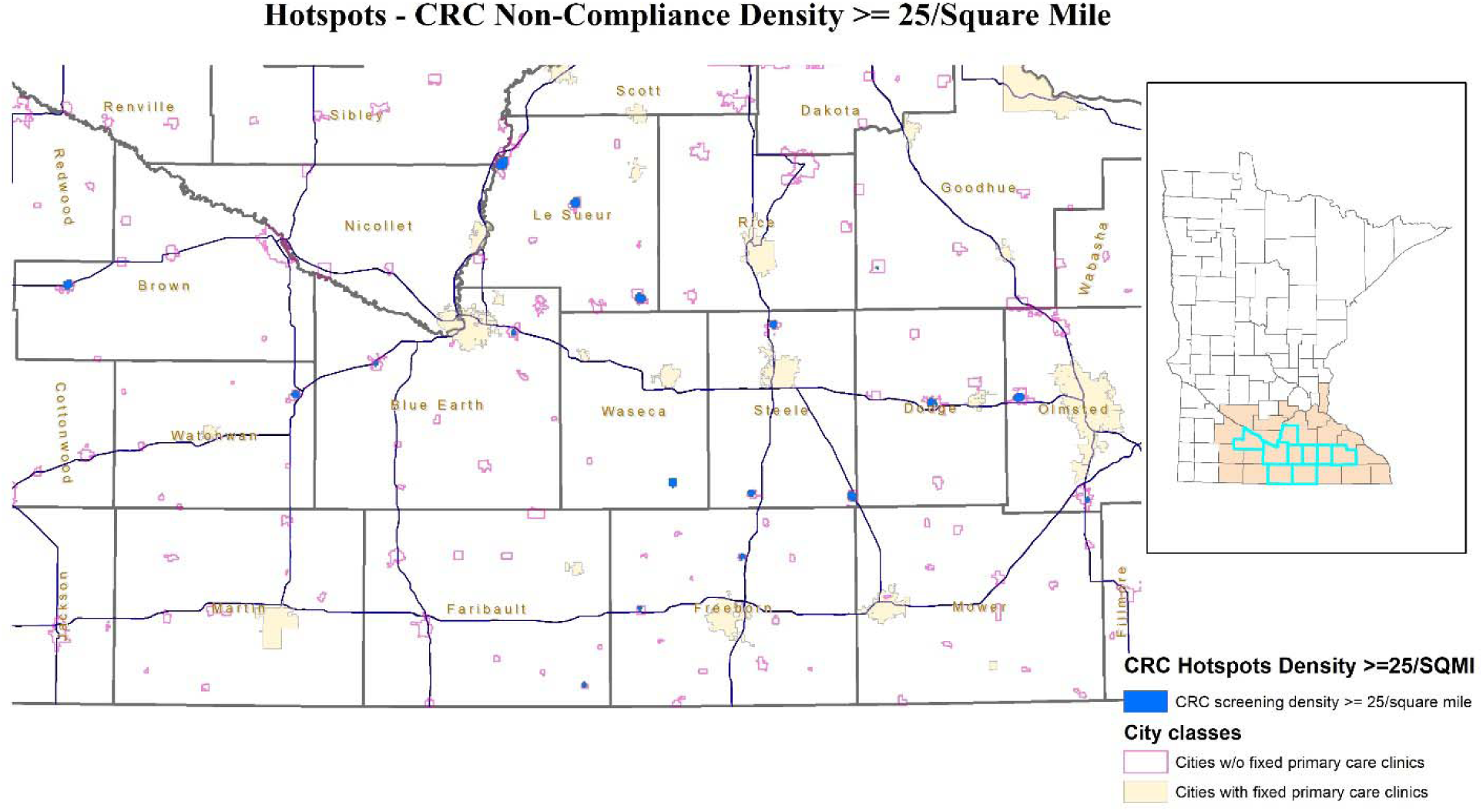
Geospatial Distribution of Low Colorectal Cancer (CRC) Screening Adherence in Rural Communities

**Figure 3.**
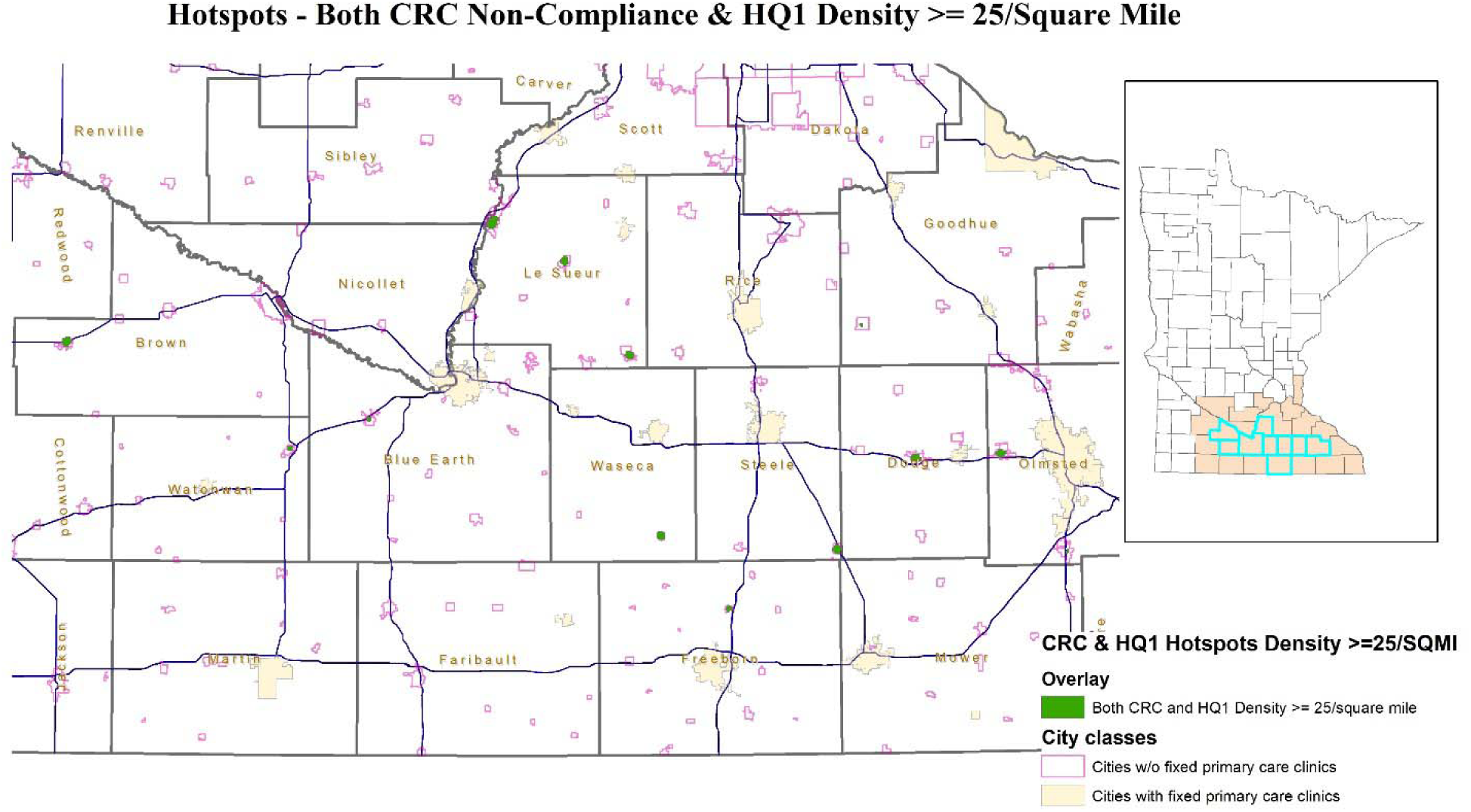
Geospatial Identification of Hotspots of Low Colorectal Cancer (CRC) Screening Adherence and Low Socioeconomic Status (HOUSES Quartile 1) in Rural Communities

One thousand two hundred twenty-one (3.5%) resided in overlapped hotspots of both lower CRC screening rates and of lower SES of the subject population (HOUSES in Q1), and 33,268 resided outside the overlapped hotspots (i.e., 1,074 (3.1%) in HOUSES Q1 only hotspot (without overlap), 633 (1.8%) CRC only hotspots, and 31,561 (91.5%) outside both hotspots). People living within the overlapped hotspots, compared to those living outside overlaid hotspots, were of lower age (63 vs 64 years old), Hispanic (3.8% vs. 0.7%), live farther from a fixed primary care clinic of MCHS (91.0 mi vs. 64.6 mi), lower CRC screening rates (69.4% vs. 79.6%), and lowest SES (HOUSES Q1 39.9% vs. 13.0%) (Table 3 and Supplementary Table 1).

**Table 3.** Baseline characteristics of study individuals in and out of both hotspots.

|  |  |  |  |  |
| --- | --- | --- | --- | --- |
|  | **Within overlapped hotspots (N=1,221)** | **Outside overlapped hotspots (N=33,268)** | **Total (N=34,489)** | **p value** |
| **CRC Screening, n (%)\*** |  |  |  | **N/A** |
| Adherent | 847 (69.4%) | 26,469 (79.6%) | 27,316 (79.2%) |  |
| Non-Adherent | 374 (30.6%) | 6,799 (20.4%) | 7,173 (20.8%) |  |
| **Age, years, median (Q1, Q3)** | 63 (58, 69) | 64 (58, 69) | 63 (58, 69) | 0.024<sup>1</sup> |
| **Female, n (%)** | 661 (54.1%) | 17,415 (52.3%) | 18,076 (52.4%) | 0.219<sup>2</sup> |
| **Race, n (%)** |  |  |  | <0.001<sup>2</sup> |
| White | 1,151 (94.3%) | 32,308 (97.2%) | 33,459 (97.1%) |  |
| American Indian/Alaska Native | 3 (0.2%) | 45 (0.1%) | 48 (0.1%) |  |
| Asian | 17 (1.4%) | 180 (0.5%) | 197 (0.6%) |  |
| Black or African American | 12 (1.0%) | 109 (0.3%) | 121 (0.4%) |  |
| Native Hawaiian/Pacific Islander | 2 (0.2%) | 13 (<0.1%) | 15 (<0.1%) |  |
| Other/Unknown | 35 (2.9%) | 598 (1.8%) | 633 (1.8%) |  |
| N-Miss | 1 | 15 | 16 |  |
| **Hispanic or Latino, n (%)** | 46 (3.8%) | 237 (0.7%) | 283 (0.8%) | <0.001<sup>2</sup> |
| **Charlson Comorbidity Index, median (Q1, Q3)** | 2.0 (1.0, 3.0) | 2.0 (0.0, 3.0) | 2.0 (0.0, 3.0) | <0.001<sup>1</sup> |
| **Distance to Clinic (mi), Median (Q1, Q3)** | 91.0 (81.1, 154.5) | 64.6 (34.5, 108.1) | 64.7 (35.5, 109.5) | <0.001<sup>1</sup> |
| **HOUSES Quartile, n (%)\*** |  |  |  | **N/A** |
| 4 (highest SES) | 114 (10.1%) | 11,617 (39.3%) | 11,731 (38.2%) |  |
| 3 | 254 (22.4%) | 8,352 (28.3%) | 8,606 (28.1%) |  |
| 2 | 313 (27.6%) | 5,727 (19.4%) | 6,040 (19.7%) |  |
| 1 | 452 (39.9%) | 3,846 (13.0%) | 4,298 (14.0%) |  |
| N-Miss | 88 | 3726 | 3814 |  |
| **Area Deprivation Index Quartile (ADI), n (%)** |  |  |  | <0.001<sup>2</sup> |
| 1 (highest SES) | 0 (0.0%) | 2,607 (8.8%) | 2,607 (8.5%) |  |
| 2 | 11 (1.0%) | 15,520 (52.6%) | 15,531 (50.7%) |  |
| 3 | 665 (58.7%) | 8,964 (30.4%) | 9,629 (31.4%) |  |
| 4 | 457 (40.3%) | 2,410 (8.2%) | 2,867 (9.4%) |  |
| N-Miss | 88 | 3767 | 3,855 |  |
| * P-value was not included for screening rates and HOUSES quartile since it is the variable that the hotspots are determined by (i.e., not independent measures); 1. Kruskal-Wallis rank sum test; 2. Pearson's Chi-squared test |  |  |  |  |

## DISCUSSION

To our knowledge, this is the first study to integrate geographic data with individual-level socioeconomic measures to identify where targeted interventions such as mobile screening units [46] may most effectively improve CRC screening adherence and reduce socioeconomic disparities in rural communities. By overlaying heatmaps of individual-level SES with screening adherence data, our study findings can generate a comprehensive view of community needs and thus guide the design of more effective interventions. This approach could further be utilized in rural health systems beyond the MCHS system by linking HOUSES data to other registry-based preventive screening records and using geospatial mapping tools to identify priority outreach areas. Smaller or non-integrated systems could implement similar approaches by partnering with public health departments to generate HOUSES-linked risk maps even when data infrastructure is limited.

Notably, our findings demonstrate that areas with both low SES (HOUSES Q1) and low CRC screening adherence showed substantial clustering. While individuals who were non-adherent to CRC screening lived farther from cities with a fixed primary care clinic of MCHS on average (Table 1), controlling for it did not find the estimate to be statistically significant (adj. OR [95% CI]: 1.00[0.998, 1.002]). These results suggest that factors beyond physical proximity influence screening behaviors, as indicated by differences in CRC screening adherence between HOUSES Q1 individuals within and outside HOUSES Q1 hotspots. This finding underscores the value of incorporating individual-level SES data into public health strategies and demonstrates the HOUSES index’s utility as a surrogate measure for identifying populations at higher risk of poor uptake of preventive services. The notable overlap between the two hotspot heatmaps further suggests the feasibility of using the HOUSES index alone for developing community outreach to address patients’ SDOH, especially when population-level CRC screening rate data is not readily available.

Addressing disparities in CRC screening is a major public health challenge, requiring interventions at individual, organizational, and population levels [47]. Ongoing efforts have focused on structural barriers, such as increasing community education and reminders [48, 49], implementing navigation programs [50, 51], and mailing non-invasive testing [52]. Our study adds to this body of work by integrating geographic data and individual-level SES, aligning with a study that employed neighborhood-level geospatial analytics to characterize screening disparities [53]. These investigations have mapped regions with the largest numbers of unscreened age-eligible individuals [54] or have examined sociodemographic factors such as SES, ethnicity, and health care facility proximity among underserved groups [7]. For instance, Zhan *et al.* used geospatial analysis and found that priorities need to be on federally qualified health centers where the number of age-eligible individuals not yet screened for CRC was the greatest [54]. Other projects have looked more broadly at mapping for different cancers with guideline-directed screening measures, such as breast and cervical cancer [29], also using variables such as the availability of cancer screening clinicians [7, 55]. However, none of the studies have integrated individual-level SES with geospatial approaches in rural areas for targeted population health management strategies for cancer screening.

In rural communities, the association between SES and CRC screening adherence is further interconnected and relevant. Financial constraints, such as transportation costs, lost income from time off work, and procedural costs, can create barriers to accessing preventive services [56]. Additionally, rural residents in low-SES areas may face challenges related to health literacy, reduced awareness of screening guidelines, and lower trust in health care providers [57, 58]. Limited access to primary care clinics and specialists, particularly in the context of increasing rural hospital closures [59], further widens these disparities [60, 61]. Lastly, psychosocial stressors and competing priorities due to economic hardship may deprioritize preventive health behaviors. All of these factors, combined with later-stage diagnoses and increased CRC mortality in rural populations, underscore the need for targeted, community-based interventions [62].

This study has limitations. First, findings from Southern Minnesota may limit generalizability to regions with different demographic or health care landscapes. However, the analytic framework may be generalizable to health care systems with similar care delivery structures and patient populations in mixed urban–rural or predominantly rural regions. Second, changes in CRC screening guidelines during the study period could have affected screening rates (e.g., increased attention and national public health campaigns). Third, we relied on the first recorded residential address in the EHR during our study time frame, which could introduce misclassification if participants relocated. An additional limitation relates to the operational definition of rurality used in this study. Rather than using a standardized rural classification alone (e.g., RUCA), we defined communities based on the absence of a fixed MCHS primary care clinic. We recognize that some of these communities may have access to health care through other health systems or independent providers. Accordingly, our definition should not be interpreted as indicating an absence of health care services overall. Instead, it was intentionally selected to reflect local MCHS service availability, consistent with the study’s implementation objective of informing targeted outreach among patients attributed to MCHS. Consequently, the findings should be interpreted as identifying communities with limited local MCHS infrastructure where outreach interventions may have the greatest operational impact, rather than as a comparison of rural versus urban populations in the traditional epidemiologic sense. Nonetheless, a notable strength of this study is the use of point data to identify concentrations, rather than on administrative boundaries (e.g., city limits) or census boundaries. In our rural areas, block groups, which were used for ADI, can be geographically large and socio-economically heterogeneous, which can obscure smaller, high-risk communities (e.g., mobile home parks). Lastly, this is a cross-sectional analysis that cannot establish causality.

Future research warrants a prospective evaluation of outcomes following targeted interventions using these heatmaps. For example, in 2021, Mayo Clinic Health System implemented a digitally equipped Mobile Health Clinic (MHC) that has efficiently served five rural communities [44]. To support the strategic expansion of this service, Mayo Clinic Health System recently launched a quality improvement initiative that used a data-driven approach, leveraging the HOUSES index and geospatial heatmap analysis, to identify additional rural communities for outreach. Effectiveness and stakeholder satisfaction associated with this targeted expansion strategy are planned in the evaluation. Additionally, assessing the scalability of these geospatial methods is essential, particularly if widespread hotspots lead to cost and logistical challenges that negate the benefits of localized resource allocation. Incorporating digital tools to increase community engagement before implementing mobile interventions could further enhance the effectiveness of these strategies. Moreover, community awareness campaigns focusing on CRC screening and the scheduling of mobile unit visits could play a vital role in improving screening uptake in underserved areas.

## CONCLUSION

Our study introduces a novel approach that integrates HOUSES index, a housing-based metric of individual-level SES, with geospatial analysis to enhance the efficacy of population health outreach initiatives in rural areas. By prioritizing high-density areas identified by heatmaps based on significant concentrations of the lowest quartile individual-level SES data, interventions such as mobile clinic deployment can more effectively allocate limited resources.

## Data Availability

The datasets generated and/or analyzed during the current study are not publicly available as they include protected health information.

## Notes

**Financial support:** This work was supported by NIH R21AG65639, NIH R01HL171508, NIH UL1 TR002377 from the National Center for Advancing Translational Sciences (NCATS), and HOUSES Program. S.B.D. was supported by the National Institutes of Health/National Institute on Minority Health and Health Disparities (NIH K23 MD016230) and Mayo Clinic Center for Clinical and Translational Science (UL1 TR02377-07).

**COI:** The authors report no relevant conflicts of interest

### Competing Interest Statement

The authors have declared no competing interest.

### Author Declarations

This study was approved by the Institutional Review Board at the Mayo Clinic (IRB # 19-009328).

