## Supplemental Table/Figure for "From Housing to Hotspots: Integrating a Housing-Based Measure of Individual Socioeconomic Status with Geospatial Analysis to Target Colorectal Cancer Screening in Rural Communities"

| **Supplementary Table 1 Baseline characteristics of study individuals overall and by type of hotspots*** | | | | | | |
| --- | --- | --- | --- | --- | --- | --- |
|  | **Overlapped hotspots (N=1,221)** | **HOUSES Q1 Only Hotspot (N=1,074)** | **CRC Only Hotspots (N=633)** | **Neither (N=31,561)** | **Total**  **(N=34,489)** | **p value** |
| **CRC Screening, n (%)*** |  |  |  |  |  | **N/A** |
| Adherent | 847 (69.4%) | 822 (76.5%) | 434 (68.6%) | 25,213 (79.9%) | 27,316 (79.2%) |  |
| Non-Adherent | 374 (30.6%) | 252 (23.5%) | 199 (31.4%) | 6,348 (20.1%) | 7,173 (20.8%) |  |
| **HOUSES Quartile, n (%)*** |  |  |  |  |  | **N/A** |
| 4 (highest SES) | 114 (10.1%) | 122 (11.6%) | 121 (27.3%) | 11,374 (40.6%) | 11,731 (38.2%) |  |
| 3 | 254 (22.4%) | 265 (25.1%) | 120 (27.0%) | 7,967 (28.4%) | 8,606 (28.1%) |  |
| 2 | 313 (27.6%) | 307 (29.1%) | 114 (25.7%) | 5,306 (18.9%) | 6,040 (19.7%) |  |
| 1 (lowest SES) | 452 (39.9%) | 361 (34.2%) | 89 (20.0%) | 3,396 (12.1%) | 4,298 (14.0%) |  |
| N-Miss | 88 | 19 | 189 | 3,518 | 3,814 |  |
| **Age, years, median (Q1, Q3)** | 63 (58, 69) | 64 (59, 70) | 64 (58, 70) | 63 (58, 69) | 63 (58, 69) | 0.012^1^ |
| **Female, n (%)** | 661 (54.1%) | 589 (54.8%) | 364 (57.5%) | 16,462 (52.2%) | 18,076 (52.4%) | 0.010^2^ |
| **Race, n (%)** |  |  |  |  |  | <0.001^2^ |
| White | 1,151 (94.3%) | 1,045 (97.3%) | 598 (94.5%) | 30,665 (97.2%) | 33,459 (97.1%) |  |
| American Indian/Alaska  Native | 3 (0.2%) | 1 (0.1%) | 0 (0.0%) | 44 (0.1%) | 48 (0.1%) |  |
| Asian | 17 (1.4%) | 3 (0.3%) | 7 (1.1%) | 170 (0.5%) | 197 (0.6%) |  |
| Black or African  American | 12 (1.0%) | 6 (0.6%) | 1 (0.2%) | 102 (0.3%) | 121 (0.4%) |  |
| Native Hawaiian/Pacific  Islander | 2 (0.2%) | 0 (0.0% | 0 (0.0%) | 13 (<0.1%) | 15 (<0.1%) |  |
| Other/Unknown | 35 (2.9%) | 19 (1.8%) | 27 (4.3%) | 552 (1.7%) | 633 (1.8%) |  |
| N-Miss | 1 | 0 | 0 | 15 | 16 |  |
| **Hispanic or Latino, n (%)** | 46 (3.8%) | 27 (2.5%) | 15 (2.4%) | 195 (0.6%) | 283 (0.8%) | <0.001^2^ |
| **Charlson Comorbidity Index,**  **median (Q1, Q3)** | 2.0 (1.0, 3.0) | 2.0 (1.0, 3.0) | 2.0 (1.0, 3.0) | 2.0 (0.0, 3.0) | 2.0 (0.0, 3.0) | <0.001^1^ |
| **Distance to Clinic (mi),**  **Median (Q1, Q3)** | 91.0 (81.1, 154.5) | 124.0 (49.3, 139.0) | 66.5 (63.4, 93.8) | 63.4 (32.6, 105.2) | 64.7 (35.5, 109.5) | <0.001^1^ |
| **Area Deprivation Index Quartile (ADI), n (%)** |  |  |  |  |  | <0.001^2^ |
| 1 (highest SES) | 0 (0.0%) | 0 (0.0%) | 0 (0.0%) | 2,607 (9.3%) | 2,607 (8.5%) |  |
| 2 | 11 (1.0%) | 133 (12.6%) | 40 (9.0%) | 15,347 (54.8%) | 15,531 (50.7%) |  |
| 3 | 665 (58.7%) | 680 (64.5%) | 311 (70.0%) | 7,973 (28.5%) | 9,629 (31.4%) |  |
| 4 | 457 (40.3%) | 242 (22.9%) | 93 (20.9%) | 2,075 (7.4%) | 2,867 (9.4%) |  |
| N-Miss | 88 | 19 | 189 | 3,559 | 3,855 |  |

* P-values were not included for screening rates since these are the variable that the hotspots are determined by (i.e., not independent measures); 1. Kruskal-Wallis rank sum test; 2. Pearson's Chi-squared test

**Supplementary Figure 1: Bar plot of counts of patients in different hotspots based on HOUSES quartile and corresponding screening status**

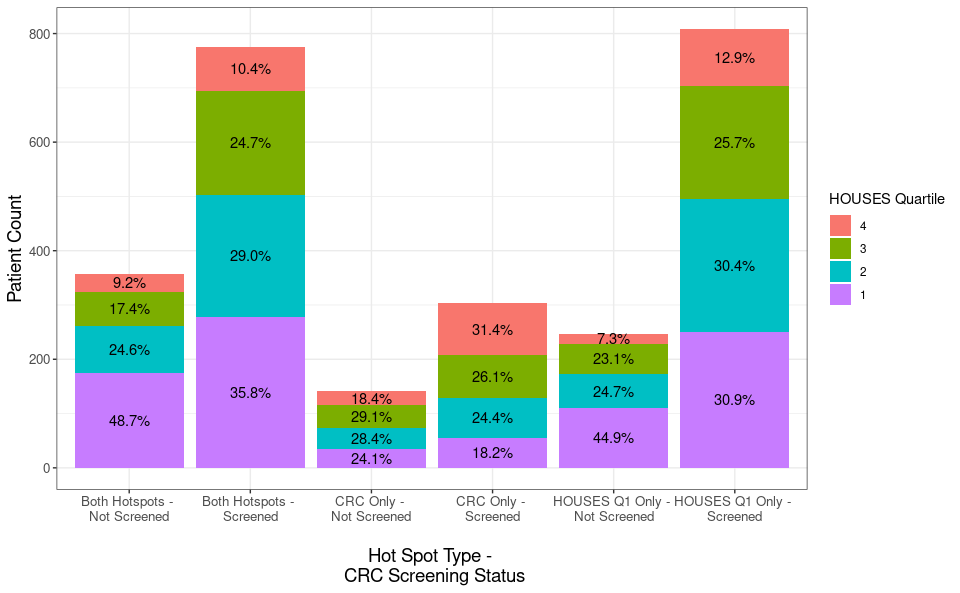
